# Mendelian randomization study of birth weight and risk of psychiatric disorders later in life

**DOI:** 10.1101/2023.10.26.23297618

**Authors:** Xiaoping Wu, Frank Geller, Dorte Helenius, Jakob Grove, Clara Albiñana, Liselotte Vogdrup Petersen, Cynthia M. Bulik, Anders D. Børglum, Thomas Werge, Bjarni J. Vilhjálmsson, Bjarke Feenstra

## Abstract

Low birth weight has been associated with a higher risk of psychiatric disorders later in life. The underlying causal mechanisms of this relationship are however not clear. In this study, we investigate whether variation in fetal growth has a direct causal effect on mental health. Using birth weight as a proxy measure for fetal growth, we first assessed associations between observed birth weight and later diagnosis of psychiatric disorders in the Danish iPSYCH and ANGI-DK cohorts. Next, we constructed a polygenic score for birth weight based on >1 million variants and tested for association with psychiatric disorders. Finally, using 86 single-nucleotide polymorphisms with robust fetal-only genetic associations with birth weight, we assessed the causal relationship of genetically mediated fetal growth and psychiatric disorders using Mendelian randomization analyses. We found that higher observed birth weight was associated with lower risk of several psychiatric disorders. Polygenic score analyses supported this pattern for attention deficit/hyperactivity disorder, where an increase of one standard deviation in the score for birth weight corresponded to an odds ratio of 0.85 (95% confidence interval 0.79-0.92, P=6.27×10^-5^). However, one- and two-sample Mendelian randomization analyses did not indicate a direct causal relationship between the birth weight of children and their risk of psychiatric disorders. In conclusion, our study does not support a direct causal effect of fetal growth (as proxied by birth weight) on the risk of psychiatric disorders later in life, suggesting that the observed association between birth weight and mental health is likely to be caused by other factors.

## Introduction

Many epidemiological studies have reported associations between extremes of birth weight and future risk of psychiatric disorders. Most findings indicate that lower birth weight is associated with a higher risk of psychiatric disorders later in life, including schizophrenia [1], depression [2], bipolar disorder [3], attention deficit/hyperactivity disorder (ADHD) [4, 5], and autism spectrum disorder (ASD) [6]. Studies for anorexia nervosa found no general correlation with birth weight [7], but showed an increased risk for girls who were small for gestational age [8]. However, it is not clear which causal factors underlie these associations. Research into fetal origins of individual differences in neurodevelopment and offspring mental health later in life has gained increasing attention [9, 10]. The Developmental Origins of Health and Disease (DOHaD) hypothesis proposes that adverse intrauterine environments result in fetal growth restriction and that developmental compensation to adverse environments “programs” fetal tissues in a manner that predisposes to specific diseases later in life [11]. The original focus of the DOHaD hypothesis was on cardiovascular diseases and health outcomes [12], but it has been expanded to also include fetal programming of neurodevelopment and mental health [9, 10]. So far, evidence in favor of DOHaD mechanisms has primarily come from observational epidemiological studies and animal experiments [13], and other theories have been proposed to explain observed epidemiological associations [14].

Figure 1 is a simplified schematic representation of different scenarios that could result in an observed epidemiological association between birth weight and mental health outcomes. The first scenario in panel (A) corresponds to a DOHaD mechanism, where a combined negative effect of environmental and maternal genetic factors leads to an adverse intrauterine environment that results in reduced fetal growth and developmental compensations that produce increased risk of offspring psychiatric disorders later in life. Panel (B) depicts a combined negative effect of fetal genetic variation and the intrauterine environment on fetal growth, which in turn has a direct causal effect on neurodevelopment and an increased risk of offspring psychiatric disorders. Panel (C) shows the situation where a set of fetal genetic variants that reduce fetal growth have pleiotropic effects that result in an increased risk of psychiatric disease. A good example is the fetal insulin hypothesis [14] that assumes certain fetal insulin genes having an effect on lower birth weight and high risk of type 2 diabetes. Finally, panel (D) represents a scenario where environmental and maternal genetic factors create an adverse intrauterine environment causing reduced fetal growth and the same factors increase the risk of offspring psychiatric disorders by mechanisms unrelated to pregnancy. This could for example be genetic variants in the maternal genome that result in reduced fetal growth through an adverse intrauterine environment and when passed on to the offspring also increases the risk of psychiatric disorders. Or it could capture environmental factors that adversely affect fetal growth and later increase the risk of offspring psychiatric disorders through exposure in the household during childhood.

**Figure 1.**
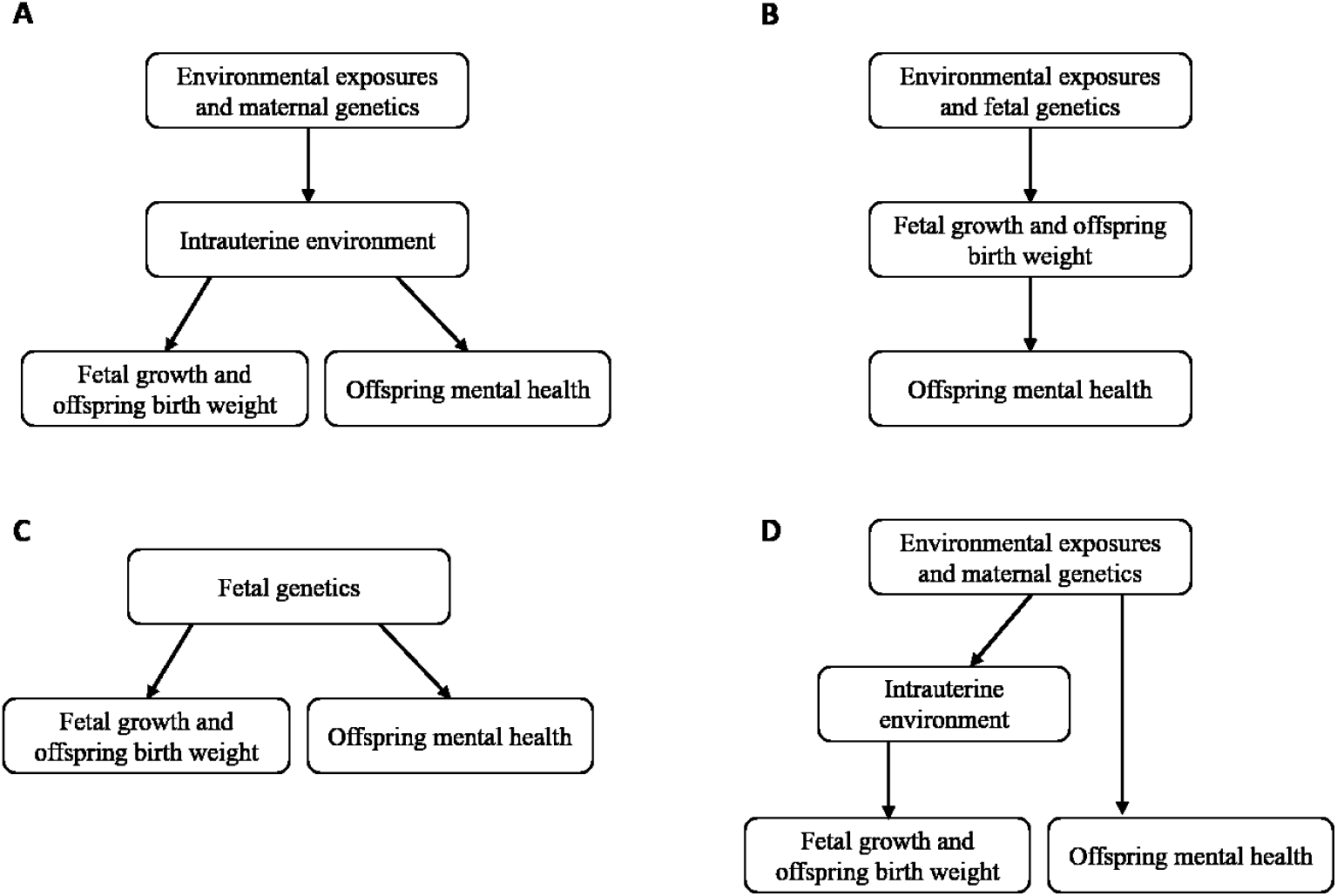
Schematic illustration of different scenarios underlying observed epidemiological associations between birth weight and mental health outcomes. **A** Environmental and maternal genetic factors in combination lead to an adverse intrauterine environment that results in reduced fetal growth and to developmental compensations that produce increased risk of offspring psychiatric disorders later in life. **B** Fetal genetic variation and the intrauterine environment have a negative effect on fetal growth, which in turn has a direct causal effect on neurodevelopment and the risk of offspring psychiatric disorders. **C** Fetal genetic variants that reduce fetal growth have pleiotropic effects that result in an increased risk of psychiatric disease. **D** Environmental and maternal genetic factors create an adverse intrauterine environment causing reduced fetal growth and the same factors increase risk of offspring psychiatric disorders by mechanisms unrelated to pregnancy.

It is challenging to draw inferences about the different scenarios and causal pathways underlying the observed associations between birth weight and offspring psychiatric disorders, since epidemiological studies can produce spurious results due to confounding, reverse causation or other sources of bias [15]. Mendelian randomization (MR) provides an alternative approach robust to most of these issues, because it utilizes genetic variants associated with an exposure of interest as instrumental variables to infer a causal effect on the outcome [15–17]. Several studies have used genetic variants associated with birth weight to instrument the intrauterine environment or fetal growth in MR studies of offspring disease outcomes later in life [18–24]. For mental health outcomes, few previous studies have been published, with conflicting results for ADHD [21, 22, 24]. In MR studies it is important to choose the right instrumental variables for the exposure. For example, a proper instrument for the intrauterine environment to test a DOHaD hypothesis (like in Figure 1A) should be based on maternal genetic variation, e.g., associated with birth weight, and ideally simultaneously controlled for fetal genotype [20]. Some previous MR studies have been problematic, e.g., using variants in the fetal genome associated with their own birth weight to instrument the intrauterine environment [25]. In this study the focus is on the scenario in Figure 1B, where fetal growth has a direct causal effect on neurodevelopment and later risk of psychiatric disorders. We are using birth weight as a proxy for fetal growth and utilize variants in the fetal genome associated with birth weight as instruments in the MR analyses.

In this work, we leverage comprehensive genotype and phenotype data from the Lundbeck Foundation Initiative for Integrative Psychiatric Research (iPSYCH2012 and iPSYCH2015i) case-cohort samples [26, 27] and the Anorexia Nervosa Genetics Initiative in Denmark (ANGI-DK) cohort [28] as well as publicly available genome-wide association study (GWAS) summary statistics to investigate the link between birth weight and six psychiatric disorders (ADHD, ASD, schizophrenia, major depression, bipolar disorder, and anorexia nervosa). Using genotype data from the two cohorts, we investigate associations between observed birth weight and the six disorders, and between polygenic scores for birth weight and the disorders. Further, using a set of SNPs that are robustly associated with birth weight (effect through the fetal genome only), we conduct one-sample MR analyses. Finally, we use GWAS summary statistics to estimate genetic correlations, and to conduct two-sample MR analyses to assess the evidence for a direct causal effect of fetal growth on offspring psychiatric disorders.

## Materials and Methods

### Subjects

The iPSYCH2012 and iPSYCH2015i are population based case–cohort samples including children born in Denmark between May 1^st^ 1981 and December 31^st^ 2008 who had a diagnosis of selected psychiatric disorders recorded in the Danish Psychiatric Central Research Register [29] or were selected as a population-based random controls [26, 27]. The ANGI-DK cohort was initiated by the Klarman Family Foundation to access individuals with history of anorexia nervosa [28] as recorded in the National Patient Register [30], and then additional genotyping was supported by the Lundbeck Foundation. The iPSYCH2012 and iPSYCH2015i samples and the ANGI-DK sample have been combined and are hereafter referred to as the iPSYCH study. In total, 141,265 iPSYCH study individuals have been genotyped, including 50,615 persons in the random population-based cohort [27].

In this study, we investigate six psychiatric disorders — ADHD (International Classification of Diseases, 10th revision (ICD-10) code: F90.0), ASD (ICD-10: F84.0, F84.1, F84.5, F84.8, or F84.9), major depression (ICD-10: F32-F39), schizophrenia (ICD-10: F20), bipolar disorder (ICD-10: F30-F31), and anorexia nervosa (ICD-10: F50.0 or F50.1). Individuals were included in the iPSYCH2012 case groups based on psychiatric diagnoses registered between 1994 and 2012; for iPSYCH2015i, psychiatric diagnoses between 1994 and 2015 were used; and for ANGI-DK diagnosis for anorexia nervosa given between 1994 and 2016 was used for inclusion. Register data was available to follow all individuals until 31 December 2016. Major depression was defined as affective disorder cases without a bipolar disorder diagnosis. The common controls used in our analyses were defined as the subset of the individuals in the population-based sample, who had no diagnosis for any of the six disorders under study recorded in the Danish Psychiatric Central Research Register. iPSYCH2012 and ANGI-DK samples were genotyped using Illumina’s PsychChip array and iPSYCH2015i samples using the Global Screening Array 2. Quality control, imputation and principal component analysis were done using Ricopili [31]. Detailed information can be seen in Grove et al. 2019 [32]. The Danish Scientific Ethics Committee, the Danish Data Protection Agency and the Danish Neonatal Screening Biobank Steering Committee approved iPSYCH2012, iPSYCH2015i and ANGI-DK, combined in the iPSYCH study.

## Statistical Methods

### Association between birth weight and psychiatric disorders

We obtained birth weight for 134,341 genotyped individuals from the Danish Medical Birth register [33]. We restricted our analyses to 125,225 individuals with gestational ages between 37 and 42 weeks as the GWAS for birth weight utilized in the MR analysis had been limited to term babies, and we excluded an additional 25,393 individuals who were not of European ancestries as accessed by a principal component analysis of the genetic data. The observed birth weights were standardized to Z-scores with mean 0 and variance 1, and three outliers were removed (standardized birth weight less than −5 or larger than 5), resulting in 99,829 individuals in the analyses (20,271 ADHD cases, 17,308 ASD cases, 4,046 schizophrenia cases, 25,969 major depression cases, 2,643 bipolar disorder cases, 4,684 anorexia nervosa cases, and 32,835 controls). Descriptive statistics for the six psychiatric disorders and the common controls are listed on Supplementary Table 1. The 32,835 control samples had not been diagnosed with any of the six disorders. Among the cases, some were diagnosed with more than one disorder (Supplementary Table 2). We performed logistic regression for the six psychiatric disorders on standardized birth weight with sex, gestational age, birth year and five principal components as covariates in the model. Birth year was included as a covariate because the case-cohorts included individuals born between 1981 and 2008. Thus, the older individuals had a higher probability of having received a diagnosis compared to the younger ones, especially for diseases with later age of onset like schizophrenia and bipolar disorder.

### Polygenic scores for fetal genetics of birth weight

We generated polygenic scores (PGS) based on fetal GWAS summary statistics for birth weight from the Early Growth Genetics (EGG) Consortium [34] using LDpred2-auto [35] and a genetic correlation matrix created from the iPSYCH cohort. The LDpred2-auto model does not need a validation data set to choose hyper-parameters [35]. The SNPs used for PGS estimation were restricted to Hapmap3 variants (https://www.sanger.ac.uk/resources/downloads/human/hapmap3.html), and we removed SNPs with MAF less than 0.01, INFO score less than 0.80, and a large discrepancy in standard error between iPSYCH cohort and the GWAS summary statistics according to the Ldpred2 recommendations [35]. A total of 1,106,129 SNPs were used to construct the PGS.

### Association between polygenic scores for fetal genetics of birth weight and six psychiatric disorders

To examine the effect of increasing PGS on birth weight and potential differences between cases and controls for each disorder, we performed linear regression of birthweight Z-scores on birth weight PGS deciles, with sex, gestational age and five principal components as covariates in the model. Deciles were chosen for a better visualization of the results. We also performed logistic regression analyses to investigate the association between PGS of birth weight and future risk of the six psychiatric disorders, including sex, gestational age, birth year, and five principal components as covariates in the model. Prior to the analyses, the PGS of birth weight were standardized to have mean 0 and variance 1.

## Mendelian Randomization

### Instrumental variables

The goal of the Mendelian randomization (MR) analyses was to investigate a potential direct causal effect of fetal growth on the risk of psychiatric disorders later in life. We therefore selected 87 SNPs as instrumental variables, which were (1) associated with birth weight at genome-wide significance in a recent GWAS [36], and (2) classified as having an effect only through the fetal genome by using model-based clustering [36]. In the iPSYCH study, the rare variant rs56188432 was missing, resulting in genotype data for 86 SNPs in the analyses. For SNPs missing in the GWAS summary statistics for the psychiatric disorders, we looked for proxy SNPs in linkage disequilibrium (r^2^>0.8) based on 1000 Genomes Phase 3 data using the LDlinkR package [37]. The proxy SNP with the highest r^2^ was selected. In the two sample MR analyses, the instrumental variables included 75 SNPs for ADHD, 83 SNPs for schizophrenia, 82 SNPs for ASD, 83 SNPs for major depression, 83 SNPs for bipolar, and 79 SNPs for anorexia nervosa. Details are given in Supplementary Table 3.

### One-sample Mendelian randomization

Using the individual-level data from the iPSYCH study, we performed one-sample MR by using the two-stage least-squares (2SLS) method [38]. First, we performed least-squares regression of the exposure (birth weight) on the 86 instrumental SNPs including sex, gestational age, birth year, five principal components, and genotype batch as covariates. Then, we performed least-squares regression of the outcome (each psychiatric disorder) on the predicted values from the first regression with the same covariates. We estimated the F statistic [39] measuring the instrumental variable strength in the 32,835 controls.

### Two-sample Mendelian randomization

The one-sample MR analyses were conducted within the iPSYCH study and thereby limited by the sample sizes available for the different disorders. Therefore, we also performed two-sample MR based on the largest GWAS meta-analyses available for the six psychiatric disorders. Details of the GWAS meta-analyses for ASD [32], ADHD [40], schizophrenia [41], major depression [42], bipolar [43] and anorexia nervosa [44] as well as for the fetal GWAS of birth weight [36] are provided in Supplementary Table 4. In all instances we used GWAS summary statistics based on individuals of European ancestries.

We first estimated the genetic correlations between birth weight and the six psychiatric disorders by bivariate linkage disequilibrium (LD) score regression [45, 46].

We estimated the statistical power of the two-sample MR analysis using the online tool by Burgess (https://sb452.shinyapps.io/power/), assuming 2% of variance explained by the instrumental variable, an odds ratio of 1.15 per SD change in exposure at an alpha of 0.05 and with the sample size of the respective study.

We conducted two-sample MR analysis using the multiplicative random-effects inverse-variance weighted (IVW) method [47, 48]. We used Cochran’s Q statistic [49] to test for heterogeneity between variant-specific causal estimates. If evidence for heterogeneity was found, we repeated the IVW analyses with a leave-one-out approach to identify whether the association was driven by a single SNP. Then, we used three additional methods for sensitivity analyses: the weighted median-based method [50], the MR-Egger method [17], and the MR-PRESSO method [51]. These methods can provide valid causal inferences under weaker assumptions: the median-based method assumes that more than 50% of the variants are valid instruments; the MR-Egger method allows all genetic variants to have pleiotropic effects; the MR-PRESSO method tests for outliers showing horizontal pleiotropy effects and provides estimates excluding these outliers.

The two-sample MR analyses were conducted with the R packages MendelianRandomization version 0.6.0 (https://cran.r-project.org/web/packages/MendelianRandomization/index.html) and MR-PRESSO [51] (https://github.com/rondolab/MR-PRESSO).

### Sensitivity analysis

To investigate the impact of maternal smoking during pregnancy and maternal mental illness on offspring psychiatric disorders, we conducted sensitivity analyses. Maternal smoking information was collected from the medical birth register (available since 1991), and maternal mental illness diagnoses (defined by ICD-10: F00-F99 and corresponding ICD-8: 290-315) were collected from the Danish Psychiatric Central Research Register and the Danish National Patient Register. Using the logistic regression model as described before, we included sex, gestational age, five principal components, as well as maternal smoking and / or maternal mental illness as covariates to investigate if maternal smoking and / or mental illness have an effect on the association between observed birth weight / PGS of birth weight and the six psychiatric disorders.

## Results

We performed logistic regression analyses of associations between observed birth weight or PGS of birth weight based on fetal genetic associations and risk of psychiatric disorders, and then used MR analyses to investigate potential causal relationships as illustrated in Figure 2.

**Figure 2.**
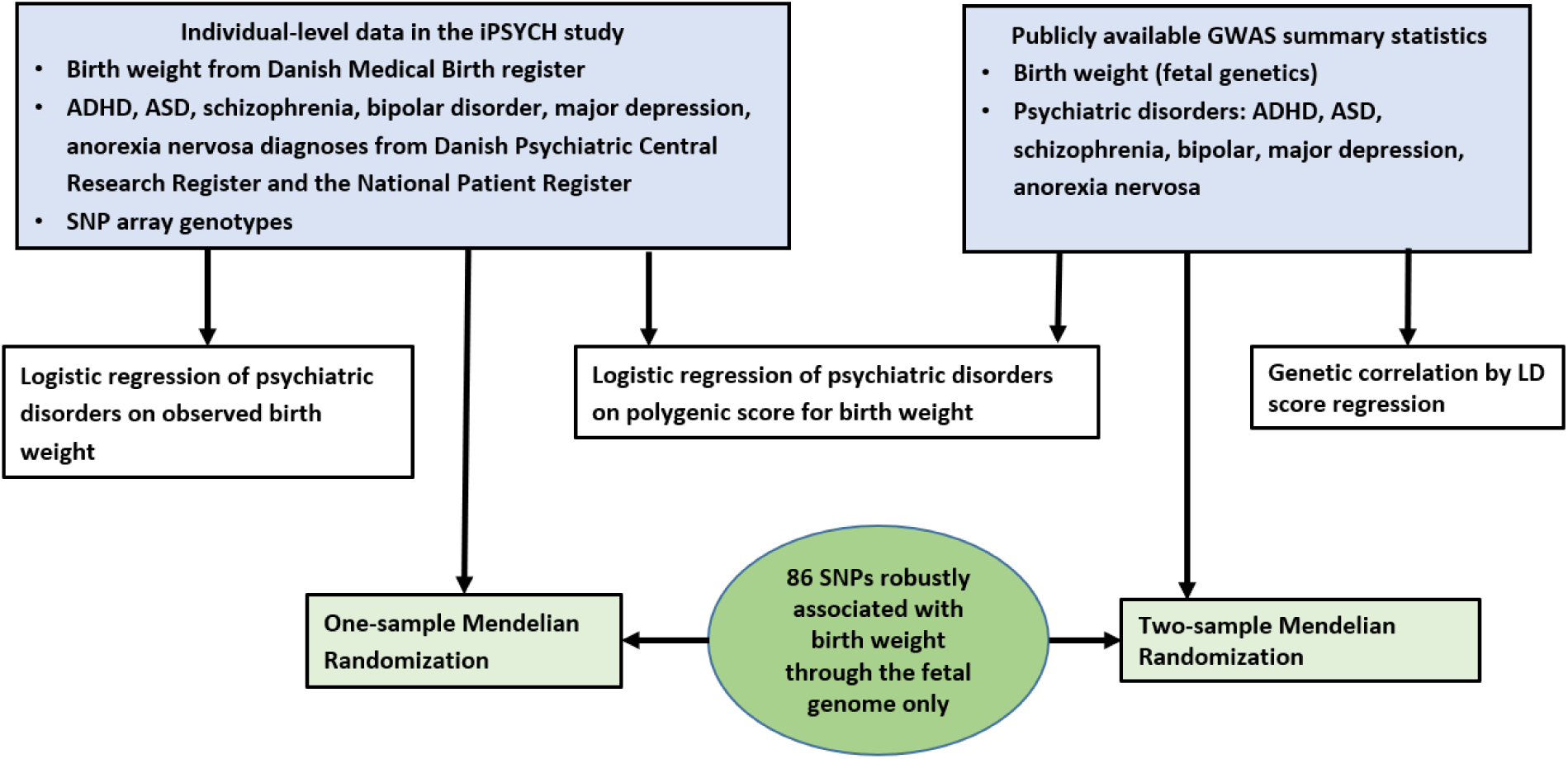
Overall study design.

### Association between observed birth weight and psychiatric disorders

In line with other studies, we found that increased observed birth weight for individuals in the iPSYCH cohort was associated with reduced risk of several psychiatric disorders. As seen in Figure 3A, an increase by one standard deviation of the Z-score of observed birth weight was associated with reduced risk of ADHD (OR = 0.87, [95% CI 0.85-0.89], P =8.08×10^-44^), schizophrenia (OR = 0.92, [95% CI 0.88-0.95], P =9.36×10^-6^), ASD (OR = 0.94, [95% CI 0.92-0.96], P =1.98×10^-8^), and major depression (OR = 0.96, [95% CI 0.94-0.98], P =1.68×10^-5^).

**Figure 3.**
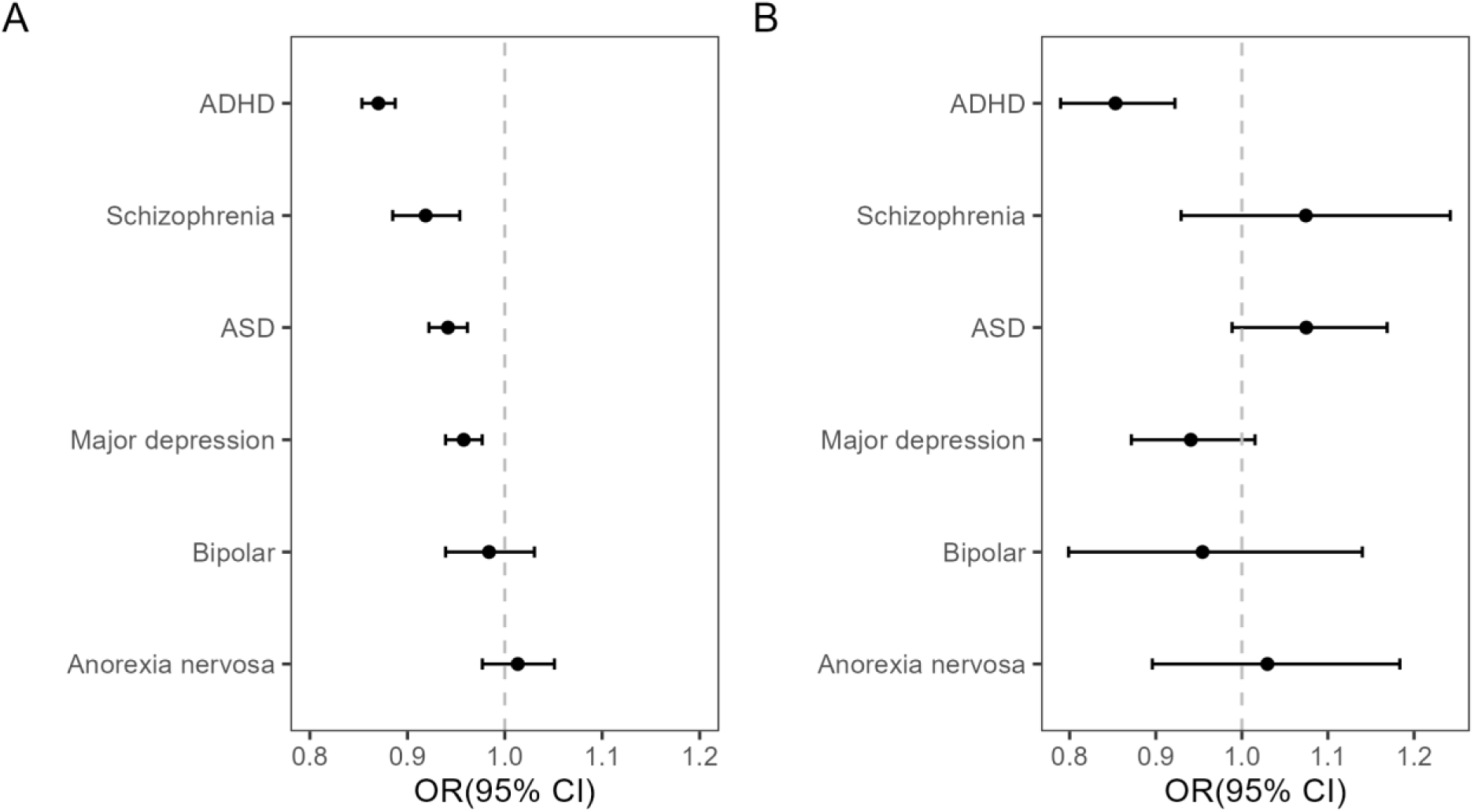
Logistic regression of psychiatric disorders in the iPSYCH study on Z-score of observed birth weight (A) and polygenic score for birth weight (B). Abbreviations: CI, confidence interval; ASD, autism spectrum; ADHD, attention deficit/hyperactivity disorder. The associations displayed in panels A and B were adjusted by sex, gestational age, birth year and five principal components and show the effect of an increase Z-score for birth weight (A) or polygenic score for birth weight (B) by one standard deviation on risk of each of the six disorders.

There was no association between observed birth weight and bipolar disorder (OR = 0.98, [95% CI 0.94-1.03], P =0.49) and anorexia nervosa (OR = 1.01, [95% CI 0.98-1.05], P =0.48).

### Polygenic score analyses

The PGS were constructed as a summary measure for each individual, combining birth weight associations of common fetal genetic variants across the genome. As expected, linear regression of Z-scores of observed birth weight on deciles of PGS for birth weight showed increasing patterns for the controls as well as for the six different psychiatric disorders (Supplementary Figure 1). Of note, for ADHD, the observed birth weight for a given PGS decile appeared to be lower compared to the control group. In logistic regression analyses, we found that an increase in PGS for birth weight based on fetal genetic associations showed negative association with ADHD (OR = 0.85 per PGS standard deviation, [95% CI 0.79-0.92], P =6.27×10^-5^) (Figure 3B).

### Genetic correlation between birth weight and psychiatric disorders

We performed bivariate LD score regression analyses using publicly available GWAS summary statistics. Results showed a negative genetic correlation between birth weight and ADHD (r_g_=- 0.078, SE=0.03, P=3.0×10^-4^) and no correlation with the five other psychiatric disorders (Figure 4).

**Figure 4.**
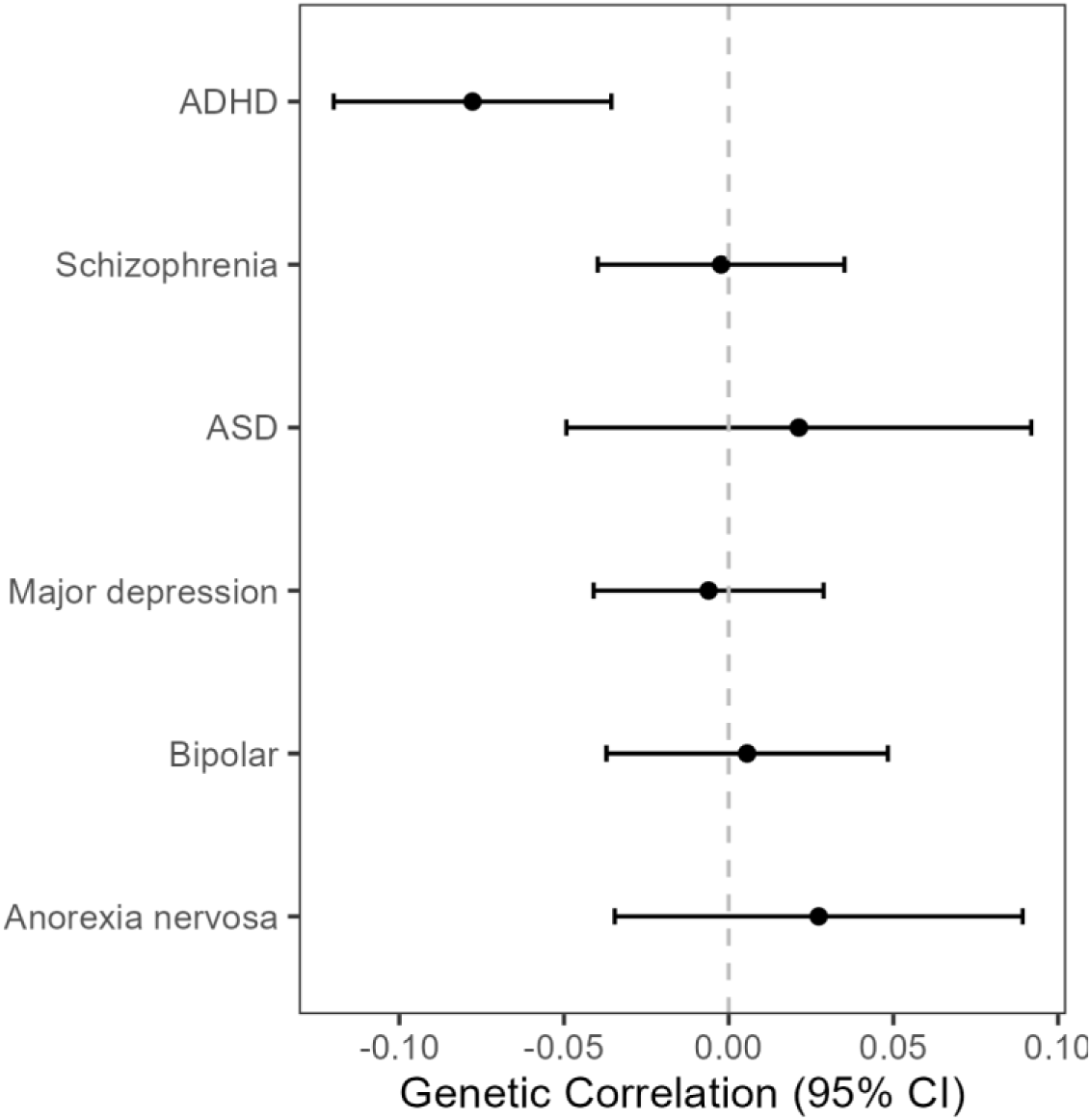
Genetic correlation between birth weight and psychiatric disorders by bivariate LD score regression. Abbreviation: CI, confidence interval; ASD, autism spectrum; ADHD, attention deficit/hyperactivity disorder.

### Mendelian randomization analyses

We assessed the potential causal relationship between fetal growth (as proxied by birth weight) and the six psychiatric disorders by MR analyses using 86 birth weight SNPs [36] as instrumental variables. These were robustly associated, genome-wide significant SNPs classified as having an effect only through the fetal genome. The analyses include: 1) one-sample MR with birth weight as exposure and each psychiatric disorder as outcome using individual-level data; 2) two-sample MR analyses with birth weight as exposure and each psychiatric disorder as outcome using publicly available GWAS summary statistics. In the two-sample MR analyses, we conducted a primary analysis (IVW method) and three sensitivity analyses (MR-Egger, weighted median and MR-PRESSO).

### One-sample Mendelian randomization analyses using individual-level data

The F statistic was 10.30, exceeding the recommended threshold of 10 for the instrumental variable to be sufficiently strong for MR analysis [52]. The analyses showed no evidence for a causal effect of birth weight on any of the six psychiatric disorders (Figure 5).

**Figure 5.**
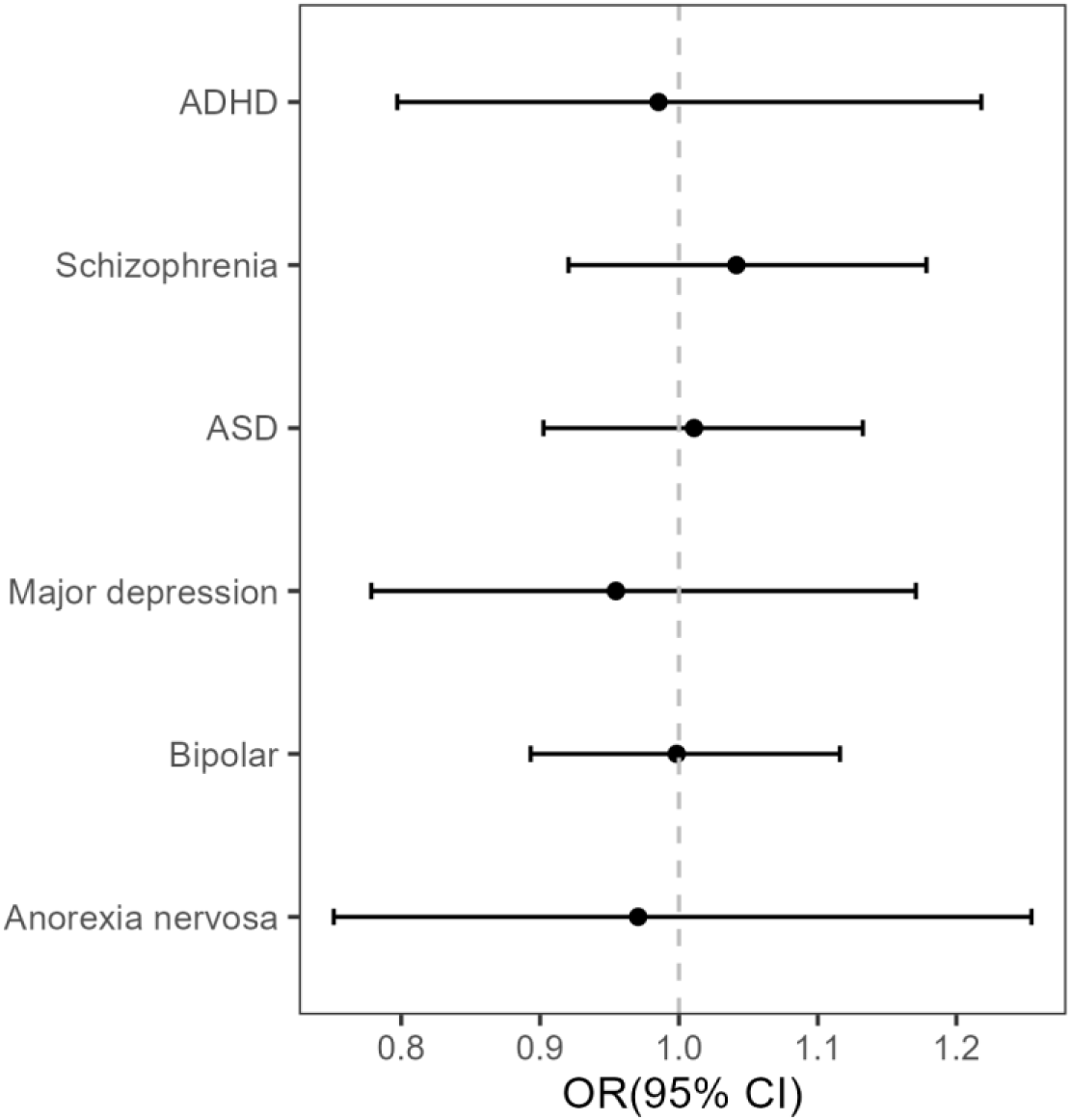
Results of the one-sample Mendelian randomization analyses testing causality in the association between birth weight and six psychiatric disorders in the iPSYCH study. Error bars represent 95% confidence intervals. Abbreviation: CI, confidence interval; ADHD, attention deficit/hyperactivity disorder; ASD, autism spectrum disorder.

### Two-sample Mendelian randomization analyses using GWAS summary statistics

We complemented the one-sample MR analyses within iPSYCH with two-sample MR analyses based on publicly available summary statistics. Details of sample size, data source and power for two-sample MR are listed in Supplementary Table 4. Our analyses had power in the range from 0.55 to more than 0.99 for an odds ratio of 1.15 for a change of one SD in the exposure. Overall, we found little evidence of a causal effect of birth weight on any of the six psychiatric disorders by two-sample MR (Figure 6), only for schizophrenia we obtained a nominally significant result with the MR-PRESSO method.

**Figure 6.**
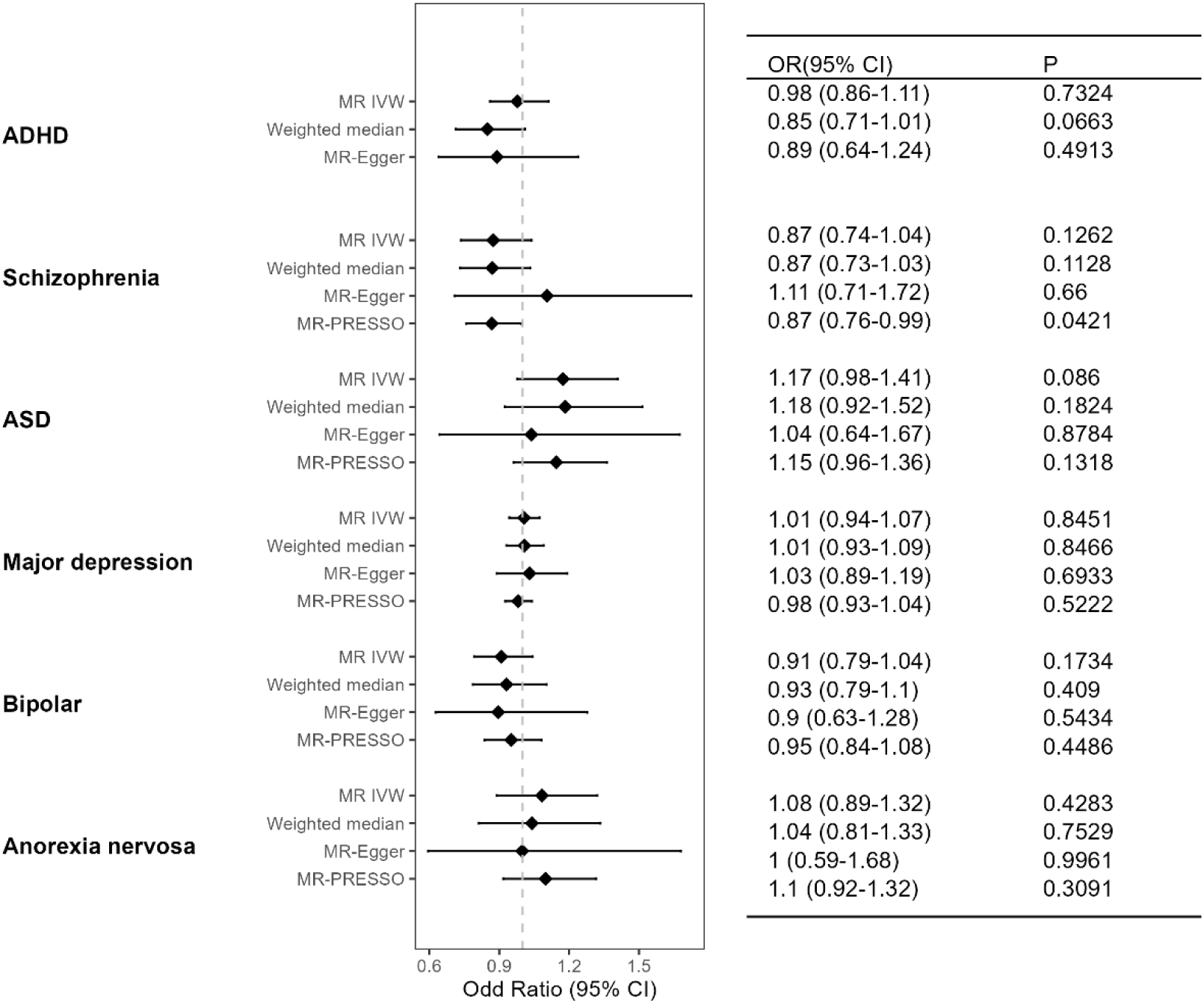
Two-sample Mendelian randomization analyses for the association between birth weight and psychiatric disorders. Error bars represent the 95% confidence intervals. Abbreviation: MR IVW, Mendelian randomization inverse-variance weighted method; ASD, autism spectrum disorder; ADHD, attention deficit/hyperactivity disorder.

Cochran’s Q statistic indicated heterogeneity between variant-specific causal estimates for all six two-sample MR analyses (Supplementary Table 4). We did leave-one-out analysis and detected one outlier SNP (rs1374204) for schizophrenia and one outlier SNP (rs7968682) for ASD. Rerunning the IVW analyses using the remaining SNPs as instrumental variables changed the estimates slightly for schizophrenia (OR=0.84, [95% CI 0.705 - 0.999], P=0.048) and ASD (OR=1.21, [95% CI 1.006 – 1.461], P=0.045). However, the sensitivity analysis by the weighted median and MR-Egger methods did not support a causal association between birth weight and these two traits.

The MR-PRESSO global test detected horizontal pleiotropy effects for all six two-sample Mendelian randomization analyses (Supplementary Table 5). Subsequent outlier tests detected between 0 and 9 outliers. However, distortion tests indicated that removing these outliers did not result in significant differences in the causal estimates (all P > 0.2).

### Sensitivity analysis

Maternal smoking during pregnancy and maternal mental illness are potential confounders in the analyses of observed birth weight or polygenic score for birth weight and psychiatric disorders (Figure 3). From the Medical Birth Register, information about smoking during pregnancy was available for 62,153 mothers, including 18,507 smokers. Among the 99,832 iPSYCH individuals analyzed there were 17,167 whose mothers had a mental illness diagnosis. As seen in Supplementary Figure 2, maternal smoking during pregnancy had a notable attenuating effect on associations between observed birth weight and psychiatric disorders. However, ADHD stood out; in analyses adjusted for both maternal smoking during pregnancy and diagnosis of maternal mental illness, we found that higher birth weight was still associated with reduced risk of ADHD (OR=0.96, [95% CI 0.94-0.98], P =7.54×10^-4^) and higher PGS of birth weight was also still associated with reduced risk of ADHD (OR=0.89, [95% CI 0.81-0.98], P =0.02).

## Discussion

Low birth weight has been linked epidemiologically to increased risk of later symptoms and diagnoses of psychiatric disorders in the child [1–6, 8, 53]. In this study, we investigated potential underlying causal mechanisms. Using Mendelian randomization analyses, we addressed the hypothetical scenario that restricted fetal growth has a direct adverse causal effect on neurodevelopment and later mental health. Our results, based on analyses of large genetic data sets did not support a direct causal effect of fetal growth on the risk of psychiatric disorders later in life, suggesting that other causal pathways are more likely to be of primary importance.

Drawing inference about causal pathways linking the intrauterine environment, fetal growth and development and later health outcomes is inherently difficult, since epidemiological studies can be subject to confounding, reverse causation, and other sources of bias [15]. Mendelian randomization is gaining popularity as an approach for causal inference in epidemiology that is robust to most of these issues. Special caution should however be exerted in MR analyses of exposures during pregnancy to properly account for the relationship between fetal and maternal genotypes, as spurious results may otherwise arise [25]. For example, if maternal SNPs associated with birth weight are used to instrument the intrauterine environment, and some of the SNPs have independent effects on birth weight through the fetal genome, a naïve MR analysis may be flawed [25]. Since we only had offspring genotypes available in our data, we focused on the scenario of a direct causal effect of fetal growth (Figure 1B). Importantly, we exclusively used SNPs that (1) were associated with birth weight at genome-wide significance, and (2) were classified as having an effect through the fetal genome only [36]. The importance of this strict approach is particularly notable when considering the results for ADHD. Here, a PGS for birth weight based on >1 million SNPs across the genome showed a clear association with risk of ADHD (Figure 3B), whereas no association was seen in one- and two-sample MR analyses based on 86 SNPs with fetal-only effects (Figures 5-6). This could suggest that the PGS is picking up effects of maternal genetic variation, corresponding to, e.g., a DOHaD scenario (Figure 1A), or to a scenario where maternal genetics influences fetal growth through the intrauterine environment and independently affects the child’s risk of ADHD through mechanisms unrelated to pregnancy (Figure 1D). The latter scenario, could for example entail mothers with a genetic predisposition to ADHD tending to smoke more during pregnancy leading to lower birth weight and at the same time passing on genetic susceptibility to ADHD to their children [54]. An ideal setup to distinguish between these different scenarios would be MR analyses in cohorts with relevant phenotypes and a large number of genotyped mother-child pairs, as has been done for other outcomes in the HUNT cohort [20].

There have been few previous MR studies of birth weight and mental health outcomes with somewhat conflicting results. Arafat et al. [21] found no causal effect of low birth weight on depression, schizophrenia, and ADHD; and Solar Artigas et al. [24] found no causal effect of birth weight on ADHD; whereas Orri et al. [22] reported a causal effect of birth weight on ADHD, post-traumatic stress disorder, and suicide attempts. In comparison to these efforts, our study had a number of strengths, including using the largest, most recent GWAS of birth weight for the classification of fetal-effect only SNPs in the instrumental variable creation [36]. Also, although previous studies have all taken a two-sample MR approach, we also conducted one-sample MR analyses with good instrument strength (*F* statistic = 10.30) in the large, well-phenotyped iPSYCH study and found consistent results between our one- and two-sample MR analyses.

Our study also had limitations. First, birth weight is an imperfect proxy measure of fetal growth and there may be aspects of fetal growth as an exposure that were not captured well in our MR analyses. In general, robust genetic associations with more refined measures of fetal growth, e.g. obtained through large-scale GWAS of ultrasound parameters at different timepoints of pregnancy would be highly warranted. Second, GWAS meta-analyses of birth weight usually exclude children born before 37 weeks of gestation [34, 36], so variants with specific effects on severely restricted fetal growth may not be captured well, as most of the very low birth weights occur in children born preterm. Third, our analyses were limited to individuals of European ancestries. Fourth, although we included both one- and two-sample MR analyses and used additional methods robust to violations of MR assumptions, it is recommended that evidence from MR studies should be triangulated with findings based on different approaches that have different sources of bias [55].

In conclusion, it is of great public health importance to understand the causal relationships that underlie associations between conditions during pregnancy and later health outcomes for the child. Our results, based on Mendelian randomization analyses of large genetic data sets, did not support a direct causal effect of fetal growth on the risk of psychiatric disorders later in life. Additional carefully designed studies are needed to investigate alternative mechanistic pathways explaining the correlation between birth weight and psychiatric disorders.

## Data Availability

All data produced in the present study (other than sensitive data) is available upon reasonable request to the authors. Owing to the sensitive nature of the iPSYCH data, individual level data can be accessed only through secure servers where download of individual level information is prohibited. Each scientific project must be approved before initiation, and approval is granted to a specific Danish research institution. International researchers may gain data access through collaboration with a Danish research institution. More information about the iPSYCH project can be obtained at https://ipsych.dk/en/about-ipsych/.

## Acknowledgments

The authors are grateful to Drs. David Hougaard, Ole Mors, Merete Nordentoft, and Preben Bo Mortensen for their long-standing work with the iPSYCH cohort, facilitating this study. Funding support was received from the Lundbeck Foundation for the Initiative for Integrative Psychiatric Research (iPSYCH). The Anorexia Nervosa Genetics Initiative (ANGI) was an initiative of the Klarman Family Foundation, and additional genotyping was supported by the Lundbeck Foundation (grant no. R276-2018-4581). The Danish National Biobank resource was supported by the Novo Nordisk Foundation. High-performance computer capacity for handling and statistical analysis of iPSYCH data on the GenomeDK HPC facility was provided by the Center for Genomics and Personalized Medicine and the Centre for Integrative Sequencing, iSEQ, Aarhus University, Denmark (grant to A.D.B.). The study was partially funded by the A.P. Møller Foundation for the Advancement of Medical Science (L-2022-00301) and the Jascha Foundation (2023-0397) and B.F. was supported by the Novo Nordisk Foundation (NNF17OC0027594).

