## Supplementary information for "Mendelian randomization study of birth weight and risk of psychiatric disorders later in life"

### Supplementary Tables

#### Supplementary Table 1. Descriptive statistics for the six psychiatric disorders and the common controls.

| Disorder | Sample size | | | Birth weight [mean (sd)], grams | | | Gestational age [mean (sd)], weeks | | | Year of birth (range) |
|---|---|---|---|---|---|---|---|---|---|---|
| | Male | Female | All | Male | Female | All | Male | Female | All | |
| **ADHD** | 14225 | 6046 | 20271 | 3571 (531) | 3423 (507) | 3526 (528) | 39.7 (1.3) | 39.7 (1.3) | 39.7 (1.3) | 1981-2008 |
| **Schizophrenia** | 2202 | 1844 | 4046 | 3540 (518) | 3434 (493) | 3492 (509) | 39.8 (1.2) | 39.8 (1.2) | 39.8 (1.2) | 1981-2004 |
| **ASD** | 13071 | 4237 | 17308 | 3620 (536) | 3479 (516) | 3586 (535) | 39.8 (1.3) | 39.8 (1.3) | 39.8 (1.3) | 1981-2008 |
| **Major depression** | 8459 | 17510 | 25969 | 3569 (512) | 3433 (488) | 3477 (500) | 39.8 (1.2) | 39.8 (1.2) | 39.8 (1.2) | 1981-2005 |
| **Bipolar** | 974 | 1669 | 2643 | 3575 (479) | 3443 (478) | 3491 (483) | 39.8 (1.2) | 39.9 (1.2) | 39.9 (1.2) | 1981-2003 |
| **Anorexia nervosa** | 331 | 4353 | 4684 | 3610 (512) | 3477 (480) | 3487 (484) | 39.7 (1.2) | 39.8 (1.2) | 39.8 (1.2) | 1981-2008 |
| **Control** | 16668 | 16167 | 32835 | 3628 (498) | 3496 (482) | 3563 (495) | 39.8 (1.3) | 39.8 (1.3) | 39.8 (1.3) | 1981-2008 |

Abbreviations: ASD, autism spectrum; ADHD, attention deficit/hyperactivity disorder.

#### Supplementary Table 2. Number of overlapping individuals among 6 psychiatric disorders.

| Disorders | ADHD | Schizophrenia | ASD | Major depression | Bipolar disorder | Anorexia nervosa |
|---|---|---|---|---|---|---|
| **ADHD** | 20271 | 424 | 3606 | 2093 | 349 | 165 |
| **Schizophrenia** | 424 | 4046 | 338 | 1229 | 178 | 113 |
| **ASD** | 3606 | 338 | 17308 | 1810 | 128 | 219 |
| **Major depression** | 2093 | 1229 | 1810 | 25969 | 0 | 1190 |
| **Bipolar disorder** | 349 | 178 | 128 | 0 | 2643 | 69 |
| **Anorexia nervosa** | 165 | 113 | 219 | 1190 | 69 | 4684 |

Abbreviations: ASD, autism spectrum; ADHD, attention deficit/hyperactivity disorder.

**Supplementary Table 3. List of the 86 SNPs used as instrumental variables.** The table lists the SNPs with their alleles and the observed allele frequencies in the iPSYCH study and indicates the available SNPs or proxy SNPs in the psychiatric disorder GWAS summary statistics.

| SNP | CHR | POS | a1 | a2 | Allele frequency | ASD | ADHD | Schizo-phrenia | Major depression | Bipolar disorder | Anorexia nervosa* |
|---|---|---|---|---|---|---|---|---|---|---|---|
| rs1011476 | 11 | 2299035 | G | T | 0.703 | YES | YES | YES | YES | YES | YES |
| rs1012167 | 20 | 39159119 | T | C | 0.625 | YES | YES | YES | YES | YES | YES |
| rs10173538 | 2 | 160569276 | C | T | 0.600 | YES | YES | YES | YES | YES | YES |
| rs10181515 | 2 | 227019461 | C | T | 0.764 | YES | YES | YES | YES | YES | YES |
| rs10265057 | 7 | 47275737 | A | G | 0.913 | YES | YES | YES | YES | YES | NO |
| rs10414727 | 19 | 54205961 | T | A | 0.687 | YES | YES | YES | YES | YES | YES |
| rs10495563 | 2 | 9662210 | G | A | 0.350 | YES | YES | YES | YES | YES | YES |
| rs10913200 | 1 | 176521655 | G | A | 0.972 | YES | NO | YES | YES | YES | YES |
| rs11042596 | 11 | 2118860 | T | G | 0.347 | YES | rs11042594 | YES | YES | YES | YES |
| rs11055030 | 12 | 12878349 | G | C | 0.742 | YES | YES | YES | YES | YES | YES |
| rs11082304 | 18 | 20720973 | G | T | 0.493 | YES | YES | YES | YES | YES | YES |
| rs1135856 | 22 | 46440605 | T | C | 0.715 | YES | NO | YES | YES | YES | YES |
| rs116964396 | 8 | 41505849 | C | A | 0.964 | YES | YES | YES | YES | YES | YES |
| rs11867479 | 17 | 68090207 | C | T | 0.646 | YES | YES | YES | YES | YES | YES |
| rs1203876 | 20 | 22540915 | A | C | 0.950 | YES | YES | YES | YES | YES | YES |
| rs12401656 | 1 | 43456767 | G | A | 0.880 | YES | YES | YES | YES | YES | YES |
| rs12584892 | 13 | 73624534 | C | T | 0.822 | YES | YES | YES | YES | YES | YES |
| rs12656216 | 5 | 36160668 | G | A | 0.240 | YES | YES | YES | YES | YES | YES |
| rs1323438 | 9 | 119115531 | T | C | 0.293 | YES | YES | YES | YES | YES | YES |
| rs13257363 | 8 | 142252580 | G | A | 0.597 | YES | YES | YES | YES | YES | YES |
| rs13322435 | 3 | 156795468 | A | G | 0.577 | YES | YES | YES | YES | YES | YES |
| rs1374204 | 2 | 46484205 | C | T | 0.297 | YES | YES | YES | NO | YES | YES |
| rs138715366 | 7 | 44246271 | C | T | 0.990 | NO | NO | NO | YES | NO | NO |

| | | | | | | | | | | | |
|---|---|---|---|---|---|---|---|---|---|---|---|
| rs141845046 | 1 | 154987704 | C | T | 0.971 | YES | YES | YES | rs146564277 | YES | YES |
| rs145775785 | 12 | 65902265 | C | T | 0.978 | YES | NO | YES | YES | YES | NO |
| rs149567808 | 7 | 43637797 | C | T | 0.994 | NO | NO | NO | YES | NO | NO |
| rs2131354 | 4 | 145599908 | G | A | 0.465 | YES | YES | YES | YES | YES | YES |
| rs220193 | 21 | 43581308 | A | G | 0.235 | YES | YES | YES | YES | YES | YES |
| rs222857 | 17 | 7164563 | C | T | 0.421 | YES | YES | YES | YES | YES | YES |
| rs2282978 | 7 | 92264410 | T | C | 0.661 | YES | YES | YES | YES | YES | YES |
| rs2296528 | 20 | 57274151 | G | C | 0.365 | YES | YES | YES | YES | YES | YES |
| rs231848 | 11 | 2735557 | A | G | 0.437 | YES | NO | YES | YES | YES | YES |
| rs234864 | 11 | 2857297 | G | A | 0.441 | YES | YES | YES | YES | YES | YES |
| rs2529415 | 7 | 50736662 | C | T | 0.347 | YES | YES | YES | YES | YES | YES |
| rs28378473 | 9 | 139245460 | T | C | 0.265 | YES | NO | YES | YES | YES | YES |
| rs28407694 | 8 | 66861738 | C | T | 0.655 | YES | YES | YES | YES | YES | YES |
| rs28457693 | 9 | 98217348 | A | G | 0.872 | YES | YES | YES | YES | YES | YES |
| rs28681372 | 22 | 50351977 | G | A | 0.404 | YES | NO | YES | YES | YES | NO |
| rs2871865 | 15 | 99194896 | C | G | 0.902 | YES | YES | YES | YES | YES | YES |
| rs2901307 | 10 | 124128443 | C | T | 0.526 | YES | YES | YES | YES | YES | YES |
| rs2908279 | 7 | 44174857 | T | G | 0.499 | YES | YES | YES | YES | YES | YES |
| rs2928148 | 15 | 41401550 | G | A | 0.459 | YES | YES | YES | YES | YES | YES |
| rs2930973 | 11 | 69445969 | G | A | 0.497 | YES | YES | YES | YES | YES | YES |
| rs2934844 | 6 | 166142456 | A | T | 0.307 | YES | YES | YES | YES | YES | YES |
| rs34776209 | 7 | 23513093 | C | T | 0.773 | YES | YES | YES | YES | YES | YES |
| rs35953885 | 2 | 43608902 | G | A | 0.751 | YES | YES | YES | YES | YES | YES |
| rs3806315 | 1 | 214724668 | G | A | 0.408 | YES | YES | YES | YES | YES | YES |
| rs3933326 | 9 | 123633948 | A | G | 0.346 | YES | YES | YES | YES | YES | YES |
| rs41355649 | 19 | 33790556 | G | A | 0.951 | YES | NO | YES | YES | YES | YES |
| rs4444073 | 11 | 10331664 | A | C | 0.551 | YES | YES | YES | YES | YES | YES |

| | | | | | | | | | | | |
|---|---|---|---|---|---|---|---|---|---|---|---|
| rs4925109 | 17 | 17661802 | A | G | 0.320 | YES | YES | YES | YES | YES | YES |
| rs4953353 | 2 | 46567276 | G | T | 0.626 | YES | YES | YES | YES | YES | YES |
| rs516246 | 19 | 49206172 | C | T | 0.553 | YES | YES | YES | YES | YES | YES |
| rs55836809 | 13 | 28502874 | A | G | 0.789 | YES | YES | YES | YES | YES | YES |
| rs55958435 | 15 | 96852638 | A | G | 0.730 | YES | YES | YES | YES | YES | YES |
| rs56252704 | 13 | 33556228 | G | A | 0.844 | YES | YES | YES | YES | YES | YES |
| rs56711588 | 2 | 109201345 | C | T | 0.560 | YES | YES | YES | YES | YES | YES |
| rs5750561 | 22 | 38595260 | A | T | 0.392 | YES | YES | YES | YES | YES | YES |
| rs577204588 | 6 | 53021737 | T | C | 0.994 | NO | NO | NO | NO | NO | NO |
| rs6033062 | 20 | 11207419 | T | A | 0.545 | YES | YES | YES | YES | YES | YES |
| rs6104763 | 20 | 11500108 | G | A | 0.360 | YES | YES | YES | YES | YES | YES |
| rs6575803 | 14 | 101257755 | C | T | 0.888 | YES | YES | YES | YES | YES | YES |
| rs6930558 | 6 | 141878920 | G | T | 0.243 | YES | YES | YES | YES | YES | YES |
| rs7076938 | 10 | 115789375 | C | T | 0.255 | YES | YES | YES | YES | YES | YES |
| rs7205514 | 16 | 50271806 | T | G | 0.243 | rs35056091 | YES | YES | YES | YES | rs35056091 |
| rs72737240 | 1 | 215387455 | T | C | 0.762 | YES | YES | YES | YES | YES | YES |
| rs72813918 | 5 | 158437823 | C | T | 0.874 | YES | YES | YES | YES | YES | YES |
| rs7354849 | 1 | 232765308 | A | G | 0.534 | YES | YES | YES | YES | YES | YES |
| rs7402983 | 15 | 99193276 | A | C | 0.406 | YES | YES | YES | YES | YES | YES |
| rs74465052 | 10 | 89605427 | T | A | 0.944 | YES | YES | YES | YES | YES | YES |
| rs74942620 | 8 | 57499986 | G | C | 0.731 | YES | YES | YES | YES | YES | YES |
| rs75104038 | 6 | 34190104 | G | A | 0.939 | YES | YES | YES | YES | YES | YES |
| rs753381 | 20 | 39797465 | T | C | 0.452 | YES | YES | YES | YES | YES | YES |
| rs7541039 | 1 | 214176779 | C | T | 0.742 | YES | YES | YES | YES | YES | YES |
| rs7563664 | 2 | 158344455 | G | T | 0.886 | YES | YES | YES | YES | YES | YES |
| rs76094073 | 6 | 109288036 | C | G | 0.881 | YES | YES | YES | YES | YES | YES |
| rs76895963 | 12 | 4384844 | T | G | 0.976 | NO | NO | YES | YES | YES | NO |

| | | | | | | | | | | | |
|---|---|---|---|---|---|---|---|---|---|---|---|
| rs7744700 | 6 | 53349401 | T | A | 0.702 | YES | YES | YES | YES | YES | YES |
| rs7771453 | 6 | 35498632 | A | G | 0.781 | YES | YES | YES | YES | YES | YES |
| rs7968682 | 12 | 66371880 | G | T | 0.509 | YES | YES | YES | YES | YES | YES |
| rs80089232 | 7 | 73018816 | T | C | 0.935 | YES | YES | YES | YES | YES | YES |
| rs9366778 | 6 | 31269173 | G | A | 0.585 | YES | YES | YES | NO | YES | YES |
| rs9379832 | 6 | 26186200 | A | G | 0.740 | YES | YES | YES | YES | YES | YES |
| rs962554 | 6 | 142734204 | T | C | 0.725 | YES | YES | YES | YES | YES | YES |
| rs9851257 | 3 | 123125711 | T | A | 0.752 | YES | YES | YES | YES | YES | YES |
| rs9855896 | 3 | 14287150 | A | G | 0.798 | YES | YES | YES | YES | YES | YES |

YES: instrumental SNP exists in GWAS summary statistics, NO: instrumental SNP does not exist in GWAS summary statistics, Others: proxy SNPs have $r^2$>0.8 with the instrumental SNP. Abbreviations: ASD, autism spectrum; ADHD, attention deficit/hyperactivity disorder.

**Supplementary Table 4. Description of public GWAS summary statistics and results of heterogeneity tests.**

| Traits | No. of samples | No. cases | No. controls | Ancestry | Power (%) | Reference | Cochran's Q | Cochran's Q P value |
|---|---|---|---|---|---|---|---|---|
| **Fetal BW** | 423683 | | | European | | Juliusdottir et al. 2021(1) | | |
| **ASD** | 46351 | 18382 | 27969 | European | 55 | Grove et al 2019 (2) | 119.65 | 3.41E-03 |
| **ADHD** | 225534 | 38691 | 186843 | European | 94 | Demontis et al 2023 (3) | 109.24 | 4.85E-03 |
| **Schizophrenia** | 127906 | 52017 | 75889 | European | 94 | Trubetskoy et al. (4) | 281.06 | 1.29E-23 |
| **Bipolar** | 413466 | 41917 | 371549 | European | 97 | Mullins et al. 2021 (5) | 150.37 | 6.29E-06 |
| **Major depression** | 500199 | 170756 | 329443 | European | >99 | Howard et al. 2019 (6) | 162.52 | 3.01E-07 |
| **Anorexia nervosa** | 72517 | 16992 | 55525 | European | 62 | Watson et al. 2019 (7) | 129.15 | 1.84E-04 |

Abbreviation: BW, birth weight; ASD, autism spectrum; ADHD, attention deficit/hyperactivity disorder. The power calculations assume an odds ratio of 1.15 per SD change in exposure.

**Supplementary Table 5. MR-PRESSO analyses for fetal birth weight (exposure) and each psychiatric disorder (outcome).**

| Outcome | Global test pvalue | Outlier SNP | Distortion test pvalue |
|---|---|---|---|
| **ASD** | 0.0048 | rs962554 | 0.7350 |
| **ADHD** | 0.0050 | | |
| **Major depression** | <2e-4 | rs1135856, rs28457693 | 0.2422 |
| **Schizophrenia** | <2e-4 | rs10173538, rs1374204, rs222857, rs28681372, rs4925109, rs516246, rs55958435, rs7541039, rs962554 | 0.9354 |
| **Bipolar** | <2e-4 | rs141845046, rs28378473 | 0.2062 |
| **Anorexia nervosa** | 2e-4 | rs1012167, rs72813918 | 0.9058 |

Abbreviation: ASD, autism spectrum; ADHD, attention deficit/hyperactivity disorder.

### Supplementary Figures

**Supplementary Figure 1. Polygenic score analyses.** Shown are associations between polygenic score deciles and standardized observed birth weight in the iPSYCH study. Black dots show associations for the subset of the population representative sample used as controls in iPSYCH, and blue dots show associations for cases of the six different psychiatric disorders.

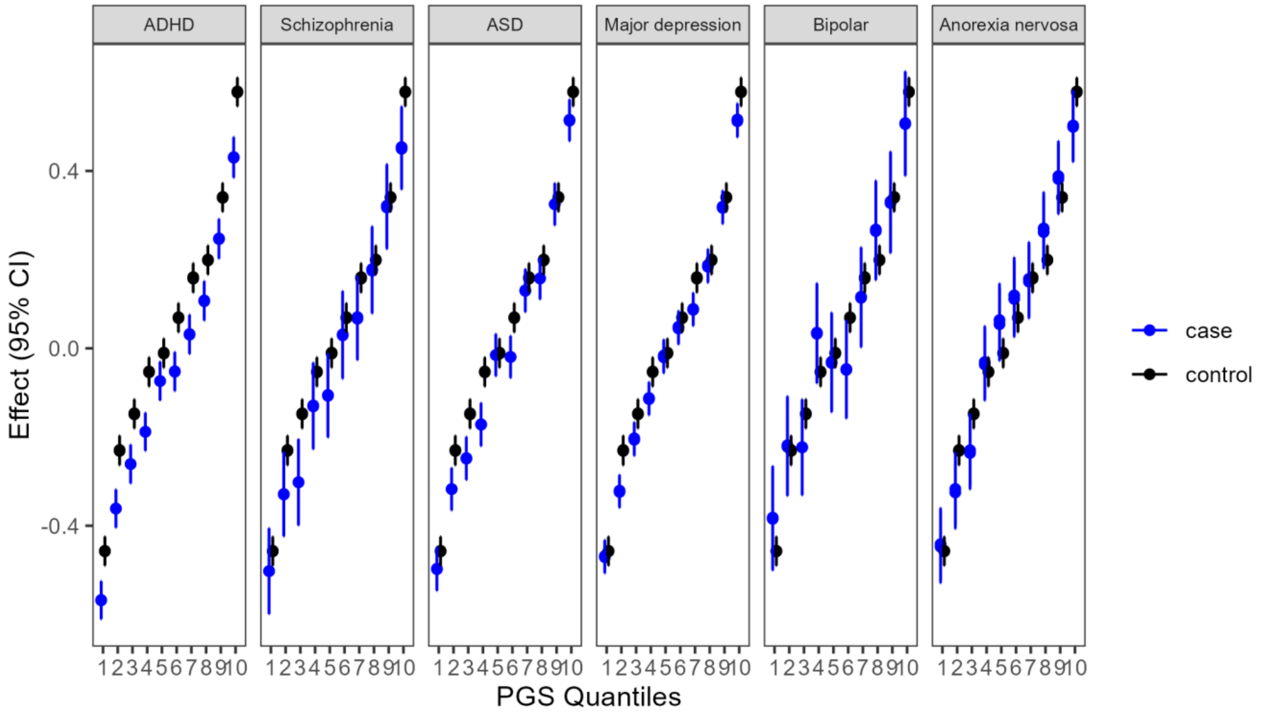


Abbreviations: CI, confidence interval; ASD, autism spectrum; ADHD, attention deficit/hyperactivity disorder.

**Supplementary Figure 2. Logistic regression of psychiatric disorders in the iPSYCH study on observed birth weight and polygenic score (PGS) of birth weight.** Analyses include maternal smoking during pregnancy, maternal mental health, and both maternal smoking during pregnancy and mental health as covariates.

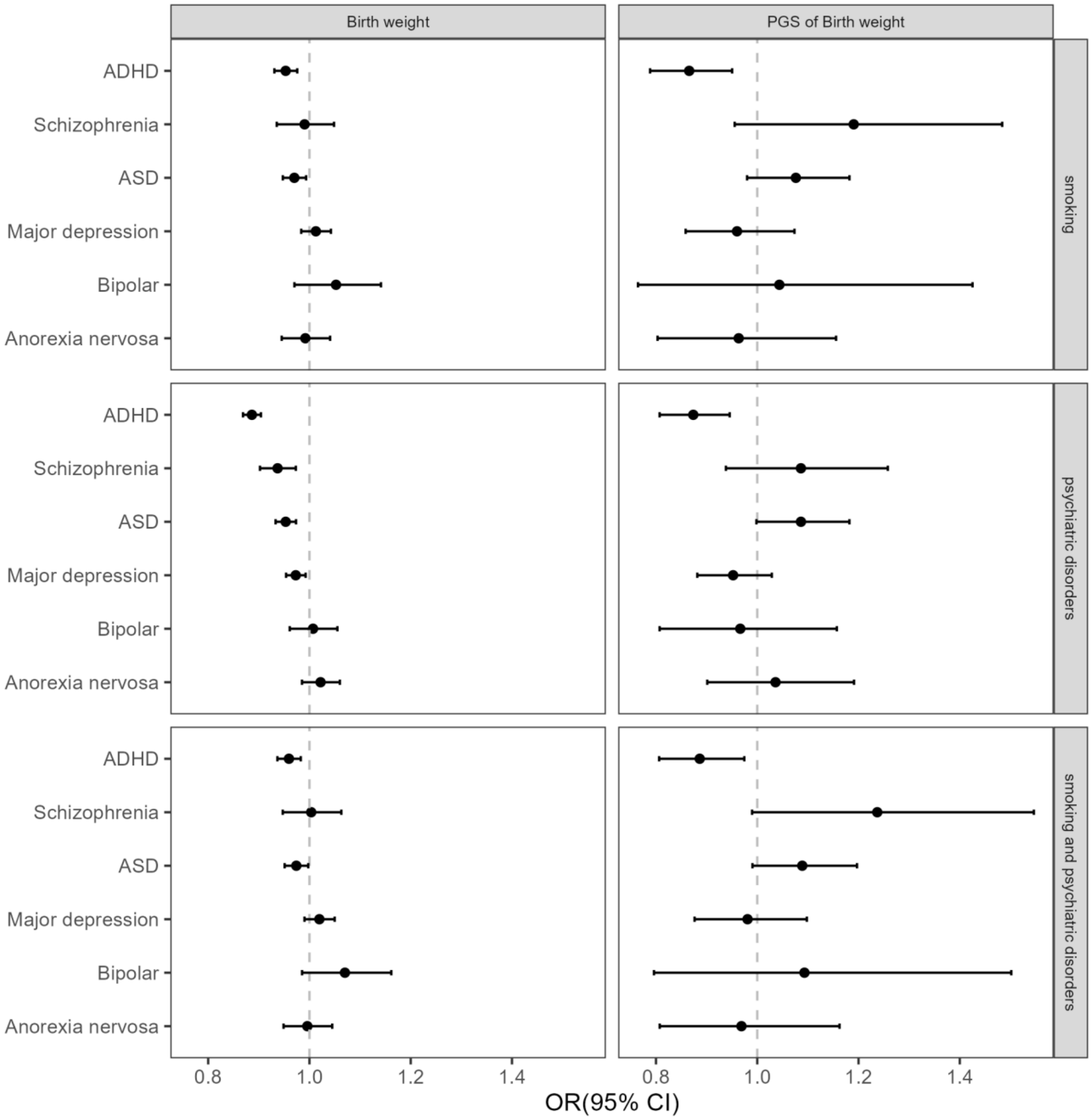


Abbreviations: CI, confidence interval; ASD, autism spectrum; ADHD, attention deficit/hyperactivity disorder.
